# An Exploratory Analysis of Essential Tremor and Associated Phenotypes

**DOI:** 10.64898/2025.12.16.25342380

**Authors:** Dylan Gharibian, Miranda Medeiros, Patrick A. Dion, Guy A. Rouleau

## Abstract

Essential Tremor (ET) is a highly heterogeneous movement disorder with a strong genetic basis. However, the etiology of ET is unclear, largely due to its clinical heterogeneity and frequently observed comorbidities.

We conducted a three-part study to investigate the genetic basis of ET in relation to co-occurring phenotypes, aiming to assess causal directionality and to clarify phenotypic heterogeneity. First, we used Mendelian Randomization (MR) to test for directional, causal relationships between ET and common co-occurring traits. We then identified pleiotropic single nucleotide polymorphisms (SNPs) shared between ET and these traits, mapped them to genes, and performed gene ontology enrichment analyses. Finally, we applied genomic structural equation modeling (g-SEM) to group traits by shared genetic variance and evaluate their influence on ET.

MR analyses did not reveal causal relationships, likely due to high genetic pleiotropy. Gene enrichment analyses of shared SNPs suggested involvement of certain pathways, but these signals were driven by limited gene overlap. SEM identified a well-fitting latent model of shared genetic architecture, but it explained only ∼2% (±9%) of ET variance.

Our findings suggest that ET and its comorbidities may share complex genetic architecture not captured by common variants alone. The limited variance explained by MR and SEM highlights the need for rare variant and multi-omics studies to better understand the biological mechanisms underlying ET and its heterogeneity.

## Introduction

Essential Tremor (ET) is among the most common movement disorders worldwide, affecting an estimated 2.87% of people over the age of 80. [1] ET is characterized by a bilateral action tremor (defined as an involuntary, rhythmic, oscillatory movement of a body part), usually affecting upper limbs, however the head and more rarely legs may be involved as well. [2] Despite the widespread prevalence of ET, [1] its etiology remains elusive. [3] This is likely due in part to the heterogeneity observed in ET, not only with regards to the nature of the tremor, [2] but also with regards to it’s numerous comorbidities. [4] Indeed, it has been suggested that the study and diagnosis of ET should be stratified by cooccurrence with psychiatric symptoms, cognitive impairment, and the presence of other movement disorders. [2] Despite the overall elusive etiology of ET, there is a strong genetic component. [3, 5] Genome Wide Association Study (GWAS) results have identified genetic loci implicated in ET, [3, 5] but a disease mechanism is still lacking, and the driving force behind the high levels of comorbidity with ET is unclear. Our study aimed to leverage GWAS data for ET as well as other associated traits to identify causal effects between them, or to find aberrant biological processes associated with both conditions. Additionally, we sought to qualify the shared genetic architecture between ET and unobserved latent variables composed of covarying traits.

The ET-associated phenotypes we chose to test in this study were informed by literature available for ET and other movement disorders. We chose to focus on areas where clear gaps were present in the literature. Psychiatric phenotypes, like bipolar disorder, major depressive disorder, and insomnia are seen to be more common in ET patients compared to healthy controls. [2] There is debate as to whether the increased prevalence of depression in ET patients is a primary feature of the disease, or a result of a reduced quality of life. [6] This highlights the need for directional studies. Cognitive impairment has been also noted with ET. [2] Environmental risk factors such as coffee consumption, alcohol intake, and smoking have been previously studied in the context of ET, however with contentious results. [7] Neurotoxic β-carbolines such as harmane, found in the diet and in coffee as well as many alcoholic beverages, have been found to be elevated in ET patients compared to healthy individuals, however it has not been shown that dietary β-carboline exposure is significantly increased in ET patients. [7] Furthermore, the role of ethanol in ET is debated, as it shows temporary relief of symptoms, but is also a known cerebellar toxin, and thus may contribute to ET pathology. [7] Smoking, which is thought to be protective in other movement disorders (like Parkinson’s disease) has also been suggested as a protective factor in ET. [8] Vitamin D is also known to be implicated in the pathology of other movement disorders, however it’s role in ET is understudied. [9] Another comorbidity of ET, hearing loss, could shed light on a shared neurological pathway relevant to both conditions. [10] Cardiological risk factors, such as hypertension and low-density-lipoprotein (LDL) levels, have also been reported to be elevated in ET patients compared to controls, however the reason remains unclear. [4] Finally, relationships between Parkinson’s disease and ET have been suggested, and the two have been shown to share genetic factors, however a shared pathological mechanism has not been found. [11]

To investigate the genetic relationship between ET and these related phenotypes, we implemented a three-part analytical strategy using SNP-phenotype associations derived from GWAS data. First, we applied Mendelian Randomization (MR) to assess potential causal relationships between ET and other traits. [12, 13] Next, we used a pleiotropy-focused approach to identify shared genetic variants between ET and individual comorbid traits, mapping these to genes and analyzing their potential convergence on shared biological pathways through gene ontology enrichment. [14] Finally, to account for potential shared underlying influences across multiple traits, we employed genomic structural equation modeling (g-SEM) to model latent genetic factors that may jointly contribute to ET. [15]

By integrating causal inference, pleiotropy analysis, and multivariate modeling, our goal was to better understand the etiology of ET and identify potential pathways or mechanisms that may underlie its clinical heterogeneity.

## Methods

### 1. Mendelian Randomization (MR)

The goal of this experiment was to measure the strength and direction of a causative correlation between ET and several other traits presumed to be related. To do this, full summary statistics were collected from the largest and most current publicly available GWASes for ET and other traits, as outlined in **Table 1**. Summary statistics from each trait were manually cleaned. Then, significant SNPs (p-value < 5×10^-8^) for each phenotype were selected as candidate instrumental variables (IVs). SNPs were harmonized using TwoSampleMR. [12, 13] Multiallelic SNPs and palindromic SNPs with intermediate allele frequencies (∼0.5) were deemed ambiguous and discarded. Steiger filtering was then carried out to remove IVs explaining a greater variance in the outcome than the exposure, as they likely effect the outcome directly (through horizontal pleiotropy). [12] MR results were calculated using the Egger Regression, [16] Inverse Variance Weighted (IVW), [17] Weighed Median (WM), [18] and Simple and Weighted modes. [19] F-statistics were calculated both per-SNP and globally for each analysis. [20] The Egger regression is a method meant to be sensitive to pleiotropy, with its y-intercept serving as an estimate for how associated the two traits are. [17] Then the global F-statistic was then calculated as described elsewhere [20]

**Table 1:** GWAS data source publications and sample sizes of tested phenotypes: *for multi-ancestry GWASes, the European ancestry cohort of the population was extracted, and reported sample sizes are from exclusively that subset.

| Trait | Publication date | Reference number | Number of cases | Number of controls |
| --- | --- | --- | --- | --- |
| Essential Tremor | 2024 | [5] | 16,480 | 1,936,173 |
| Bipolar Disorder | 2021 | [23] | 41,917 | 371,549 |
| Cigarettes per Day | 2022 | [24] | 784,353 |  |
| Coffee Consumption | 2019 | [25] | 358,093 |  |
| Cognition | 2024 | [26] | 455,496 |  |
| Alcohol Consumption (Drinks per Week) | 2022 | [24] | 2,965,643 |  |
| Essential Hypertension | 2019 | [25] | 99,665 | 289,307 |
| Hearing Loss | 2019 | [25] | 96,354 | 274,359 |
| Insomnia | 2022 | [27] | 593,724 | 1,771,286 |
| Low-Density Lipoprotein (LDL) Levels | 2021 | [28] | 1,320,016 |  |
| Major Depressive Disorder (MDD) | 2019 | [29] | 246,363 | 561,190 |
| Parkinson's Disease | 2019 | [30] | 39,275 | 1,411,006 |
| 25-OH Vitamin D Levels | 2023 | [31] | 421,867 |  |

Subsequently, a Leave-One-Out (LOO) plot was made, where the IVW score was recalculated with each IV iteratively left out, to ensure that no IV was significantly skewing the results, as this suggests confounding. [13] The Pleiotropy RESidual Sum Outlier (PRESSO) method was also used to detect outliers in the regression, based on the r^2^ values and data simulations. [21] Linear regression plots were made for each analysis, as well as forest plots. Each analysis was conducted bidirectionally. Bonferroni correction was applied to the 32 analyses (16 traits × 2 directions), giving a minimum p-value significance threshold of 0.0016. [22]

### 2. PolarMorphism and Enrichr Gene Ontologies

Summary statistics from each trait were aligned with ET, similarly as for the MR section, however without extracting only significant SNPs for a single trait. This was done using the PolarMorphism R package’s AlleleFlip function. [32] Polar coordinates for the trait-ET pairs were then calculated using the ConvertToPolar function from PolarMorphism. [32] SNPs that had a strong magnitude of effect (r > 4), intermediate angles, and a strong adjusted p-value for the angles (θ q-value < 0.05) were extracted as pleiotropic. The shared pleiotropic SNPs were then mapped to genes using the NIH SNP database (dbSNP) API. [14] The Enrichr gene ontology dataset “GO Biological Processes 2025” was then used to find molecular pathways in which shared ET-trait genes are implicated. [33-35]

### 3. Genomic Structural Equation Modelling

Using the same GWAS studies as for the MR experiment, genomic structural equation modeling was carried out to characterize the pleiotropy relationships between traits. Summary statistics were reformatted and a covariance matrix was created using linkage disequilibrium score regression (LDSC). [36] As all summary statistics were processed using effective sample sizes, sample prevalences were set to 0.5 for all binary traits (and NA for continuous traits), as is suggested by the GenomicSEM guidelines. [15] Population prevalences were manually inputted for Bipolar disorder (0.028), [37] Essential Hypertension (0.26), [38] ET (0.0032), [1] Hearing loss (0.20), [39] Insomnia (0.06), [40] MDD (0.083), [41] and Parkinson’s disease (0.00572). [42] The 1000 Genomes LD score reference file for European populations was used for the construction of the matrix. [43] An exploratory factor analysis was then run using the ‘stats’ R package and used to guide the partition of traits into various latent variables for the construction of a structural equation model. [44] Additionally, the Enrichr “GWAS Catalog 2023” dataset was used to identify phenotypes containing many overlapping genetic signals, to inform genetic sharedness for latent variable selection. [34] A handful of models were tested with different numbers of latent variables and different subsets of traits. The model with the highest Comparative Fit Index (CFI) [45] and lowest Standardized Root Mean Squared Residual (SRMR) [46] was chosen.

## Results

### 1. Mendelian Randomization

The overall objective of the mendelian randomization experiment was to infer directional association between ET and other traits, using SNP-derived instrument variables from the GWAS of each trait. No exposure trait was able to confer significant risk for ET across all analytical methods. Similarly, ET as an exposure did not have robust association with any outcome traits across analytical methods. The null hypothesis cannot be rejected for any of the trait-ET pairs in any direction (threshold p-value of 0.0016 after Bonferroni correction). These results are summarized in **Figure 1**, which presents a comprehensive forest plot of bidirectional MR results across all traits, and in **Table 2**, which lists the number of IVs retained per analysis and notes whether any MR analytical method detected a significant effect.

**Table 2:** Number of instrumental variables retained by test and significant results.

| *Exposure* | *Outcome* | *Number of IVs retained* | *Significant result(s)* |
| --- | --- | --- | --- |
| ET | Bipolar disorder | 27 | N/A |
| Bipolar | ET | 58 | N/A |
| ET | Cigarettes smoked per day | 27 | N/A |
| Cigarettes smoked per day | ET | 56 | N/A |
| ET | Coffee consumption | 26 | N/A |
| Coffee Consumption | ET | 34 | N/A |
| ET | Cognition | 21 | N/A |
| Cognition | ET | 50 | N/A |
| ET | Drinks (alcohol) consumed per week | 27 | N/A |
| Drinks (alcohol) consumed per week | ET | 83 | N/A |
| ET | Essential hypertension | 27 | N/A |
| Essential hypertension | ET | 165 | N/A |
| ET | Hearing loss | 27 | N/A |
| Hearing loss | ET | 30 | N/A |
| ET | Insomnia | 12 | N/A |
| Insomnia | ET | 7 | N/A |
| ET | LDL levels | 25 | N/A |
| LDL levels | ET | 438 | N/A |
| ET | MDD | 24 | N/A |
| MDD | ET | 32 | N/A |
| ET | PD | 25 | Inverse variance weighted<br>($p = 5.06e-03$) |
| PD | ET | 23 | N/A |
| ET | 25-OH Vitamin D | 27 | N/A |
| 25-OH Vitamin D | ET | 122 | MR Egger<br>($p = 4.72e-02$) |

**Figure 1:**
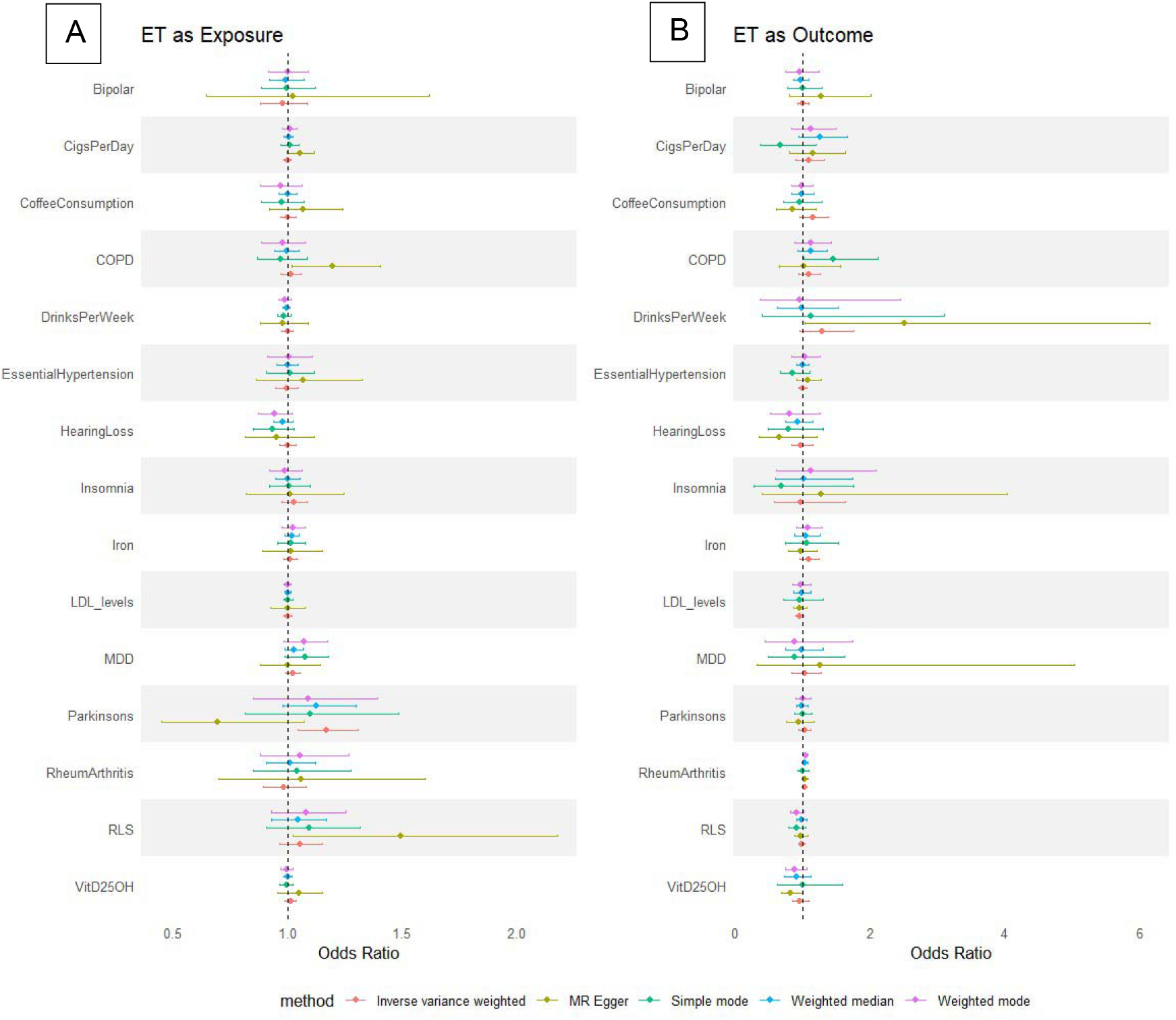
Bidirectional MR results for essential tremor with all phenotypes. The forest plots depict MR analyses results when (A) essential tremor is the exposure with all other traits as outcomes, and MR results when (B) all other traits are the exposure and essential tremor is the outcome.

While no causal or directional relationships were supported by the MR results, several analyses revealed notable features related to horizontal pleiotropy. In particular, the analysis of ET and Parkinson’s disease demonstrated a striking Egger intercept in one direction, consistent with a high degree of pleiotropy between these traits. Similarly, several analyses, such as those involving LDL levels, MDD, and alcohol intake, showed large shifts in MR effect estimates after outlier removal or Steiger filtering, further suggesting that shared genetic architecture may confound causal inference in these cases.

Detailed forest plots and sensitivity diagnostics for each trait pair, including Egger intercepts and LOO results, are presented in **Supplemental Figures S1-S12** (sensitivity analyses) and **Supplemental Figures S13-S23** (forest plots).

Once again, no causal and directional associations could be made between ET and other traits via MR. However, interesting horizontal pleiotropy was noted in many analyses, which may partially be obscuring MR, which seeks to regress out genetic correlations.

### 2. PolarMorphism and Enrichr Gene Ontologies

The objective of this experiment was to identify SNPs having a high degree of effect shared between ET pairwise with the other tested trait. This was done using PolarMorphism, [32] an R package that identifies pleiotropic SNPs, and Enrichr Gene ontologies, [35] to map genes containing those SNPs to biomolecular pathways.

Among all trait pairs analyzed, the strongest result was found between ET and bipolar disorder, for which PolarMorphism identified 616 pleiotropic SNPs mapping to 86 unique genes. The top ten terms most significantly enriched across biological pathways described in the Enrichr “GO Biological Processes 2025” catalogue is shown in **Supplemental Figure S24**. The most significantly enriched pathway shared between ET and bipolar disorder identified was the “Regulation of Potassium Ion Transport” term (adjusted p = 0.0126), with an odds ratio (OR) of 28.5 and a gene overlap count of four.

Another significant enrichment was observed between ET and hearing loss, with 166 shared SNPs across 36 genes. One pathway, “Anterograde Dendritic Transport,” reached statistical significance (p_adjusted_ = 0.018), with an OR of 234.8. However, this enrichment was based on just two overlapping genes out of a total of seven in the pathway, raising concerns that the result may be driven by the small size of the gene set.

Between ET and LDL cholesterol, 413 pleiotropic SNPs mapped to 62 unique genes. Two pathways (“Positive Regulation of Transcription by RNA Polymerase II” and “Regulation of Vascular Associated Smooth Muscle Cell Apoptotic Process”) had adjusted p-values of 0.0217 and 0.0370, respectively. Both results were marginal and potentially misleading: the first term is very broad (983 genes), and the second was driven by only two overlapping genes.

All other trait pairs yielded no statistically significant enrichment results after correction for multiple testing. This meant no significant pathway enrichment for genes relevant to ET and cigarettes smoked per day (203 pleiotropic SNPs mapping to 56 genes), or coffee consumption (1,816 pleiotropic SNPs mapping to 158 genes), or insomnia (1,381 pleiotropic SNPs mapped to 88 genes), or cognition (3,055 pleiotropic SNPs mapping to 318 genes), or alcohol intake (500 pleiotropic SNPs mapping to 83 genes), or essential hypertension (237 pleiotropic SNPs mapping to 42 genes), or MDD (531 pleiotropic SNPs mapping to 112 genes), or vitamin D levels (68 pleiotropic SNPs mapping to 17 genes). There was also no significant association for ET and Parkinson’s disease, though notably only 3 pleiotropic SNPs were identified, mapping to just 2 genes. Although Mendelian randomization suggested shared architecture, the paucity of SNPs in this analysis precluded meaningful pathway interpretation.

Overall, while some results did have significant adjusted p-values, upon closer inspection none hold up to rigor, with the possible exception of “Regulation of Potassium Ion Transport” in the analysis between ET and bipolar disorder. This result, however, is also more dubious because bipolar disorder is far less associated with ET than other psychiatric phenotypes, like MDD.

#### Genomic SEM

The objective of this experiment was to model the pleiotropy between ET and other traits, by creating a structural equation model linking traits to ET through overarching latent variables. This was done using the GenomicSEM R package. [15] Partitioning of traits into latent variable was informed by exploratory factor analysis, the correlation matrix (shown in **Supplemental Figure S35**) and by Enrichr GWAS Catalog results. The final model is shown in **Figure 2**.

**Figure 2:**
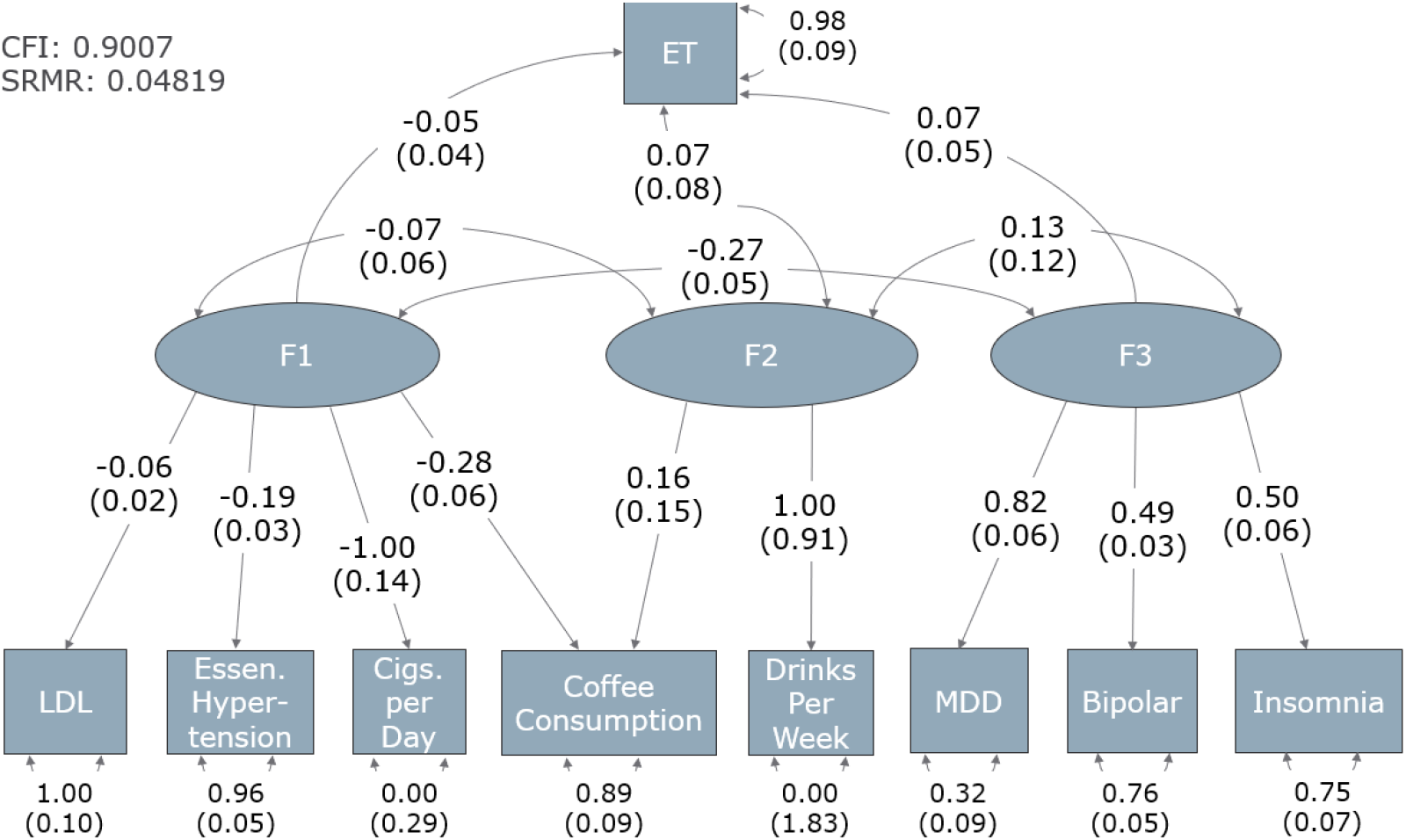
Structural Equation Model of ET and associated traits. The figure depicts the interactions between unobserved latent variables (shown in ellipses) and observed phenotypes (in squares). Unidirectional arrows from latent variables to traits represent the loading of the phenotype onto the factor, with loading coefficients written on the arrow, and standard deviations in parentheses underneath. The portion of variance in the trait unexplained by its loading on to the factor is found on the double-barbed curved arrows pointing from the trait to itself, with standard deviations in parenthesis. Finally, the curved arrows between latent variables represent the degree of interaction between latent variables, with standard deviations in parenthesis. The model metrics (CFI and SRMR) are shown in the top left corner.

In the covariance matrix (**Supplemental Figure S35**), ET is seen to have little genetic correlation with other traits, except MDD (which has been previously reported in the GWAS used in our experiment). [5] Other well-known associations are also noticed, like between bipolar disorder and MDD. From this covariance matrix, we observe that bipolar disorder, MDD, and insomnia have shared genetic architecture, therefore informing a candidate latent variable. Alcohol intake (drinks per week) and coffee consumption also share some correlation. Additionally, in examining the Enrichr results for GWAS associations from the gene lists compiled in section 2 between ET and drinks per week, and between ET and coffee consumption, it was seen both pointed to each other’s GWAS. Thus, we hypothesize that they will affect ET in a group, as a latent variable. A similar observation was made with LDL levels, smoking, and hypertension. Smoking and coffee consumption also have correlation, as seen in **Supplemental Figure S35**, and load onto the same latent variable. The final constructed structural equation model is shown below, in **Figure 2**.

Both significance metrics point to the model being significant, however the actual information given by the model is less informative. Looking at the model, we see that the amount of variance unexplained by loading onto any of the three factors for ET is 0.98, with a standard error of 0.09. Thus, most of the variance in ET cannot be explained by our analysis. Furthermore, across all factors, ET is the trait with the lowest loading, in fact being less than the amount of intercorrelation between traits. Thus, we cannot partition the pathophysiology of ET into phenotypically distinct genetic architectures. Furthermore, the high degree of uncertainty in the drinks per week trait both in the loading onto F2 and especially in the degree of variance unexplained by that loading are concerning. This is likely a result of the fact that it is the only trait loading uniquely onto F2, as both ET and coffee consumption load onto other factors as well.

## Discussion

Despite the high prevalece of essential tremor (ET), [1] its etiology remains poorly understood, with no single causative pathway, gene or exposure being identified. What is known is that ET has many comorbidities, and has a heterogenous clinical presentation. [2] Therefore, we sought to study the pathophysiology of ET together with these phenotypes, in an attempt to control for the heterogeneity in the disease and explore the relationship between ET and its associated phenotypes.

The first step was to assess whether ET has a direct effect on other phenotypes or *vice versa*. This was done using mendelian randomization (MR). [12, 13] Ultimately, we could not determine significant causal or directional relationships between ET and any other traits by MR, but interesting results were observed, nonetheless. Results bordering significance in some analytical methods prompt further study, such as the Egger regression suggesting alcohol intake as an exposure for ET. While our results are not significant, the effect of alcohol intake has long been debated in ET, as it temporarily relieves symptoms, but is also a cerebellar toxin. [7] Additionally, β-carbolines, like those found in coffee and many alcoholic beverages, as well as caffeine, have been suggested as exposures for ET. [7] While different analytic methods give contradictory results in our study of the effect of coffee consumption on ET, it is interesting to note that SNP rs1260326 (a *GCKR* missense variant) is seen by leave-one-out analysis (LOO) to be driving a positive association in both cases. The same effect driven by rs1260326 is noted with 25-OH vitamin D levels. The effect of *GCKR* (or another gene represented in the LD block of rs1260326) in mediating the effect environmental factors have on ET pathogenesis may therefore merit further study. The Egger regression of 25-OH vitamin D levels as an exposure for ET also bordered on significant in the study, suggesting (inconclusively) a mildly protective effect of vitamin D on ET. This is fascinating, as vitamin D deficiency is implicated in other movement disorders, like Huntington’s disease, restless leg syndrome, and Parkinson’s disease. [9] The large degree of pleiotropy observed between ET and PD provides further evidence for a shared genetic background between ET and other movement disorders. [2] In addition to PD, sensitivity analyses suggest that horizontal pleiotropy plays a wide-spread role in the etiology of ET, being noted with coffee consumption, alcohol intake, and MDD as well. A limitation of MR is that it is contingent on a low degree of pleiotropy between traits. [21] Additionally, MR only captures liability explained by common SNPs, which only represent an estimated 18% of ET heritability. [3]

Seeing a shared genetic architecture between ET and other phenotypes, we elected to explore the biological processes that could be genetically affected in both ET and associated traits. To do this we used PolarMorphism [32] to identify genes shared between traits, and Enrichr Gene Ontologies [33, 34] to find biomolecular processes associated with these genes. This method has several limitations, as the direction of SNP effect is ignored, as well as the actual effect the SNP would have on the gene product. Additionally, intergenic SNPs that could alter gene expression and be responsible for pathogenicity are ignored. Despite these limitations, preliminary associations of certain genes to ET in the context of specific comorbidities could still be made. Overall, even results with significant adjusted p-values were not particularly promising, either being biased by the small number of genes in certain pathways or implicating pathways that were too broad to be of use.

While examining Enrichr GWAS results, it was noted that genes were often found to be significantly associated with several phenotypes across the GWASes tested. This led us to hypothesize that examining the genetic architecture of ET may require analyzing ET with groups of comorbidities, rather than individually. To do this we employed genomic structural equation modeling (g-SEM). [15] A high-confidence model was found, and factors did make intuitive sense, as we saw psychiatric traits in one factor, lifestyle associated traits in another, and β-carboline intake associated traits (alcohol and coffee consumption) on latent variables together. Despite this, ET heritability was poorly explained by loading on to these latent variables, with 98% (with a standard deviation of 9%) of its variance remaining unexplained. Additionally, latent variables influenced each other to a greater degree than they influenced ET, so we could not alleviate heterogeneity by partitioning ET into genetically distinct subtypes. Therefore, despite the clinical observation that ET and these traits are related, we failed to find a basis to subdivide ET cases into distinct genetic subtypes.

Overall, our results failed to identify directional, causal association between ET and its associated phenotypes. We also failed to identify robust biological process associations shared in the pathogenicity of ET as well as other traits. Our g-SEM failed to partition ET into genetically distinct phenotypic pathways. Yet, a clearly established increase in frequency of certain comorbid phenotypes is present, and remains unexplained. [47] Our study suggests that we may have exhausted the capability for GWAS data to explain the relationship between ET and those comorbidities. Importantly, GWAS-targeted common variants only explain roughly 18% of ET’s heritability, [3] thus our results suggest that rare-variant and multi-omics studies across large well-phenotyped cohorts are needed to explore the syndromic nature of ET etiology.

## Data Availability

All data produced in the present study are available upon reasonable request to the authors

https://journals.plos.org/plosgenetics/article?id=10.1371/journal.pgen.1011033

https://www.nature.com/articles/s41586-022-05477-4

https://www.nature.com/articles/s41586-021-04064-3#data-availability

https://atlas.ctglab.nl/

https://www.nature.com/articles/s41593-018-0326-7#Sec2

https://www.nature.com/articles/s41588-021-00990-0

https://www.nature.com/articles/s42003-024-06207-4#Sec31

## Acknowledgements

We are thankful to the members of the Rouleau lab for their support in this work.

